# Heart Rate as a Predictor of Mortality in Heart Failure Patients at the Time of Discharge from the Intensive Care Unit

**DOI:** 10.1101/2023.07.24.23293084

**Authors:** Chia-Ying Hsiao, Min-I Su, Yu-Cheng Chang, Ying-Hsiang Lee, Po-Lin Lin, Wei-Ru Chiou

## Abstract

**Objective:** Heart rate serves as a critical prognostic factor in heart failure patients. We hypothesize that elevated heart rate in critically ill heart failure patients upon discharge from the intensive care unit (ICU) could be linked to adverse outcomes.

**Design:** We implemented a retrospective cohort study using data collected between 2008 and 2019 from the Medical Information Mart for Intensive Care IV (MIMIC-IV version 2.0) database. We examined the association between the last heart rate prior to ICU discharge and in-hospital mortality, total mortality, and ICU readmission.

**Setting:** ICU at Beth Israel Deaconess Medical Center

**Patients:** Adult patients admitted to the ICU diagnosed with heart failure.

**Interventions:** None

**Measurements and Main Results:** From the 76,943 ICU stays, we enrolled 2,365 patients in this study. We observed correlations between in-hospital mortality and ICU discharge heart rate of 83.56±15.81 bpm (survivors) vs. 93.84±17.28 bpm (nonsurvivors, p<0.001). Total mortality showed similar trends, with 83.67±15.36 bpm (survivors) vs. 85.23±17.25 bpm (nonsurvivors, p=0.027), as did ICU readmissions at 83.55±15.77 bpm (nonreadmitted) vs. 88.64±17.49 bpm (readmitted, p<0.001). Given multivariate analysis, the ICU discharge heart rate strongly predicted in-hospital mortality (OR 1.035 [95% CI 1.024-1.046], p < 0.001), total mortality (OR 1.007 [95% CI 1.001-1.014], p = 0.027) and ICU readmission (OR 1.015 [95% CI 1.007-1.023], p < 0.001). Patients with an ICU discharge heart rate >90 bpm demonstrated significantly higher in-hospital mortality (OR 2.986 [95% CI 2.066-4.315], p < 0.001), total mortality (OR 1.341 [95% CI 1.083-1.661], p = 0.007), and ICU readmission rates (OR 1.638 [95% CI 1.270-2.114], p < 0.001).

**Conclusions:** The findings suggest that heart failure patients with an elevated heart rate (>90 bpm) at ICU discharge are more likely to experience increased in-hospital mortality, total mortality, and ICU readmissions, indicating potential negative outcomes.

**Key Points:** Question: Does an elevated heart rate at ICU discharge increase in-hospital mortality, total mortality, and ICU readmission?

Findings: This retrospective cohort study using the Medical Information Mart for Intensive Care IV database showed that a higher ICU discharge heart rate >90 bpm is a strong predictor of increased in-hospital mortality (OR 2.986, p < 0.001), total mortality (OR 1.341, p = 0.007), and ICU readmission (OR 1.638, p < 0.001).

Meaning: Heart failure patients with an elevated heart rate (>90bpm) at ICU discharge are more likely to face increased risks of in-hospital mortality, total mortality, and ICU readmissions.

## Introduction

Heart rate is related to disease and stress, and the maximum heart rate is an indicator of short-term mortality in critical illness (1). A heart rate ≥90 beats per minute (bpm) at the time of multiple organ dysfunction diagnosis is an independent risk factor for increased 28-day mortality (1). Patients presenting with prolonged elevated heart rate, defined as a heart rate >95 bpm for >12 hours in at least one 24-hour period of their intensive care unit (ICU) stay, had an increased incidence of major cardiac events among critically ill, cardiac high-risk patients and a significantly longer ICU stay (2).

Heart failure (HF) is an important public health problem (3). The sudden intensification of symptoms and signs of HF, which typically results in hospitalization, is defined as acute decompensated heart failure (ADHF) (4). According to the Romanian Acute Heart Failure Syndromes registry, 10.7% of patients with the condition required ICU care, with admission to the ICU being linked to an increased risk of in-hospital mortality (17.3% vs. 6.5%, p=0.002) (5). Within a Taiwan national database of 192,201 patients who were admitted to the ICU, a total of 25,263 (13.14%) patients were readmitted. HF was identified as a significant risk factor for readmission to the ICU (HR 3.365 [95% CI 3.047–3.717], p ≤0.001) (6). Although the level of stability required for long-term HF treatment is based on a clinical evaluation that is influenced by various factors, heart rate is one of these factors.

Higher heart rate was related to the development of regional and global LV dysfunction independent of subclinical atherosclerosis and coronary heart disease (7). There are 2 clinical risk prediction models designed to risk stratify hospitalized HF patients, including the risk factor for heart rate. They are the OPTIMIZELHF (Organized Program to Initiate Lifesaving Treatment in Hospitalized Patients With Heart Failure) (8), and the GWTGLHF (Get With The Guidelines L Heart Failure) registry (9). In OPTIMIZELHF, the mean heart rate of HF patients was 87 ± 21.5 bpm in patients surviving during the hospital stay and 89 ± 22.7 bpm in patients dying during the hospital stay. On multivariable analysis, an increase of 10 bpm in heart rate, between 65 and 110, is associated with an increase in in-hospital mortality. (OR 1.094 [95% CI 1.062L1.127], p < 0.0001). In GWTGLHF, the likelihood of in-hospital mortality can be calculated for each patient by adding up points assigned to each predictor variable for a total score ranging from 0 to 100. Heart rate is one of these factors; a rate of 79 bpm or less is assigned 0 points, whereas a rate of 105 bpm or more is assigned 8 points.

In 2001, the CIBIS II trial demonstrated that heart rate can not only function as an indirect marker of disease but may also directly contribute to the development of HF (10). The SHIFT (Systolic Heart failure Treatment with the If Inhibitor Ivabradine Trial) trial showed that ivabradine treatment helps systolic HF patients with sinus rhythm and a heart rate of ≥70 bpm (11). The first hospital admissions for worsening HF were decreased in the lowest heart-rate group (70 to <72 bpm) compared with the group with the highest heart rate at baseline (≥87 bpm) (12). In the placebo group, patients exhibiting the highest heart rate ≥ 87 bpm were found to be at more than twice the risk of cardiovascular death or hospital admission for worsening HF compared to patients with the lowest heart rate (70 to <72 bpm) (hazard ratio [HR] 2.34, 95% CI 1·84–2·98, p<0·0001), and the event rates rose by 3% for every bpm increase from the baseline heart rate and by 16% for every 5 bpm increase (12). Ivabradine treatment is associated with reduced risks of cardiovascular mortality, all-cause mortality, and HF rehospitalization within 1 year among patients with acute decompensated HF with reduced ejection fraction (HFrEF) in real-world populations (13). Among patients with HFrEF, ivabradine treatment had a more notable benefit on hemodynamic stability (14). Both the 2021 European Society of Cardiology (ESC) Heart Failure Guidelines and the 2022 American Heart Association/American College of Cardiology/Heart Failure Society of America (AHA/ACC/HFSA) Guideline for the Management of Heart Failure have given a Class IIa indication regarding ivabradine by specifical consideration(15, 16).

Based on the aforementioned information, we summarize the following key points. First, heart rhythm plays a crucial role as an important prognostic factor in patients with HF. Second, pharmacological management aimed at controlling the heart rhythm in hospitalized HF patients holds the potential to improve their prognosis. Third, it has been observed that over 10% of patients hospitalized with acute HF require intensive care, and they have a higher mortality rate. However, there is currently a lack of relevant studies investigating the association between post-treatment heart rhythm control in critically ill HF patients upon transfer from the ICU and their future clinical outcomes. Therefore, our objective in the present study is to address this research gap by exploring the aforementioned issues with the MIMIC IV database, thereby providing more comprehensive understanding and guidance for clinical practice.

## Materials and Methods

### Data Source

This cohort study was conducted using data from the Medical Information Mart for Intensive Care IV (MIMIC-IV version 2.0) database, which was collected between 2008 and 2019 (17, 18). MIMIC-IV is a publicly available and real-world clinical database that was maintained by the Beth Israel Deaconess Medical Center during the same period. It includes records of over 200,000 emergency department admissions and over 60,000 ICU stays. We were granted permission to use the database, and the code for extracting data can be found on GitHub (https://github.com/MIT-LCP/mimic-iv).

### Study Population

Our study included only adult patients who were admitted to the ICU due to HF and older than 18. The diagnoses of HF were determined using the International Classification of Diseases, tenth Revision (ICD-10), and the following codes were considered: I500, I501, I509, I110, I130, and I132. In cases where a patient was admitted to the ICU multiple times, only data from the first ICU stay were included. We eliminated from our analysis patients who were discharged from the ICU within 24 hours and those with missing data for vital signs such as blood pressure and heart rate within 24 hours before discharge (Figure 1).

**Figure 1.**
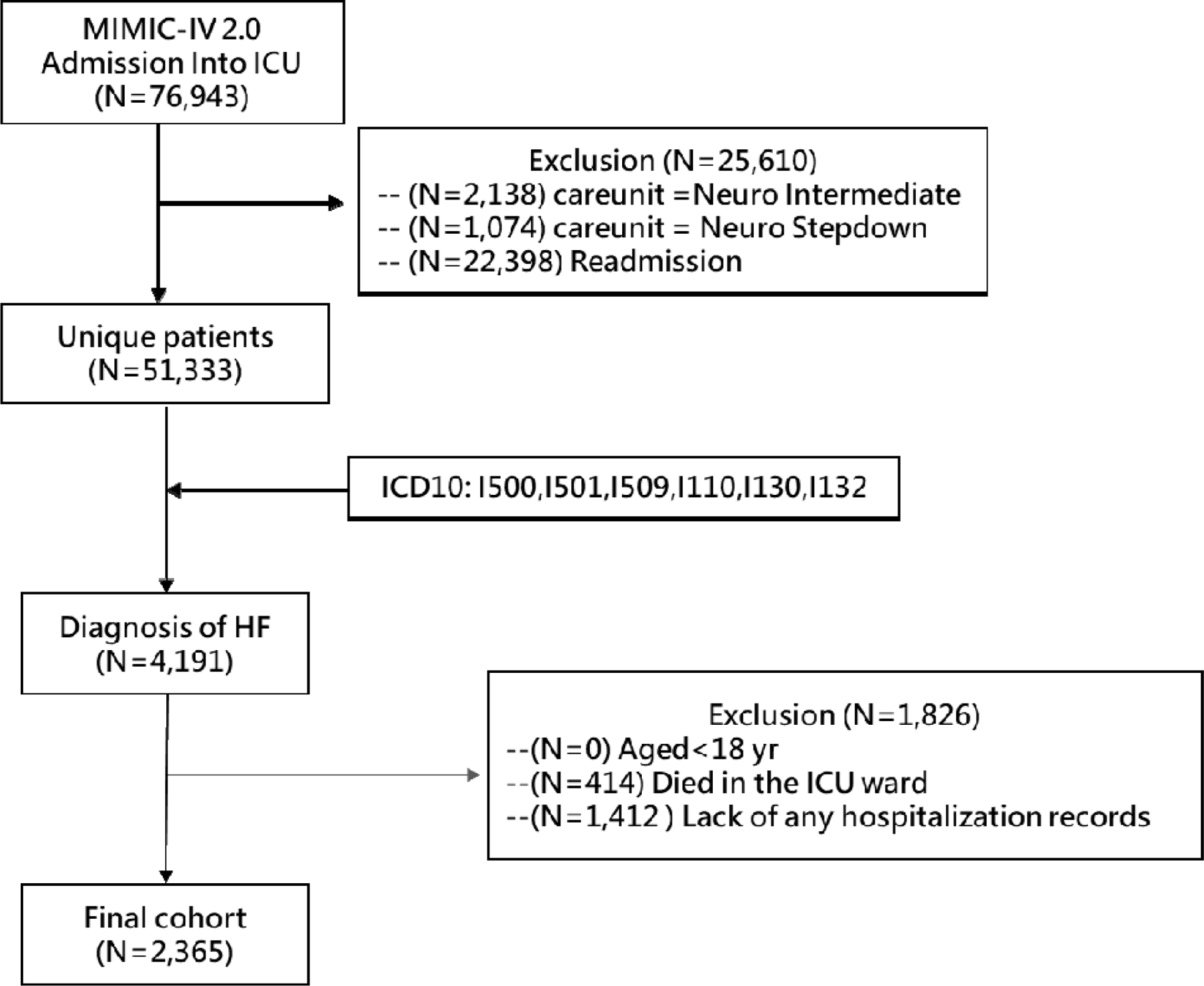
Study flowchart. *MIMIC* Medical Information Mart for Intensive Care

### Definitions and Outcomes

The ICU discharged heart rate was defined as the last heart rate before discharge from the ICU, and the average ICU discharged heart rate was defined by the 24-hour average heart rate before discharge from the ICU. The primary outcomes were in-hospital mortality and total mortality. The total mortality was populated from state death records. The secondary outcome was ICU readmission in the same hospitalization. ICU readmission was defined as when a patient was readmitted to the ICU after an interval of more than eight hours since the previous discharge from the ICU.

### Covariates

In our study, we included demographic and clinical characteristics recorded in the ICU. We also calculated various scores within the first 24 hours of ICU admission and vital signs such as heart rate, blood pressure, respiratory rate, SpO2, urine output, and serum creatinine were extracted throughout the entire ICU stay (Table 1). These covariates, which included basic demographic information and clinical characteristics of patients, were chosen based on previous related studies.

**Table 1.** Baseline characteristics between the total population and primary/secondary outcomes.

| Variable | In-hospital mortality |  | *p* | Total mortality |  | *p* | ICU readmission |  | *p* |
| --- | --- | --- | --- | --- | --- | --- | --- | --- | --- |
|  | Survival | Dead |  | Survival | Dead |  | No | Yes |  |
| Patient (%) | 2204 (93.20) | 161 (6.81) |  | 1472 (62.24) | 893 (37.76) |  | 2033 (86.96) | 332 (14.04) |  |
| Sex (F) (%) | 887 (40.25) | 70 (43.48) | 0.454 | 575 (39.06) | 382 (42.78) | 0.077 | 822 (40.43) | 135 (40.66) | 0.952 |
| Age (mean (SD)) | 69.31 (12.68) | 75.10 (11.61) | <0.001 | 67.41 (12.58) | 73.49 (11.93) | <0.001 | 69.76 (12.70) | 69.38 (12.64) | 0.619 |
| Weight (kg) (mean (SD)) | 85.37 (25.46) | 77.88 (22.58) | <0.001 | 88.02 (26.08) | 79.64 (23.15) | <0.001 | 85.04 (25.32) | 83.73 (25.46) | 0.382 |
| Height (cm) (mean (SD)) | 168.52 (11.00) | 167.52 (10.56) | 0.264 | 169.20 (10.90) | 167.20 (10.98) | <0.001 | 168.54 (10.95) | 167.90 (11.09) | 0.330 |
| BMI (kg/m<sup>2</sup>) (mean (SD)) | 29.98 (8.35) | 27.69 (7.40) | 0.001 | 30.66 (8.40) | 28.46 (7.98) | <0.001 | 29.86 (8.30) | 29.63 (8.42) | 0.640 |
| MBP (mmHg) (mean (SD)) | 84.11 (19.24) | 84.39 (17.92) | 0.857 | 84.43 (18.76) | 83.64 (19.78) | 0.331 | 84.05 (19.12) | 84.61 (19.39) | 0.622 |
| ICU Discharge HR (bpm) (mean (SD)) | 83.56 (15.81) | 93.84 (17.28) | <0.001 | 83.67 (15.36) | 85.23 (17.25) | 0.027 | 83.55 (15.77) | 88.64 (17.49) | <0.001 |
| ICU Discharge aHR (bpm) (mean (SD)) | 83.20 (14.29) | 92.41 (14.55) | <0.001 | 83.18 (14.00) | 84.89 (15.21) | 0.006 | 83.15 (14.42) | 87.94 (14.23) | <0.001 |
| ICU admission HR (bpm) (mean (SD)) | 87.13 (20.27) | 94.90 (21.66) | <0.001 | 86.86 (20.16) | 88.98 (20.88) | 0.015 | 87.09 (20.21) | 91.16 (21.63) | 0.001 |
| Temperature (°C) (mean (SD)) | 36.67 (0.69) | 36.68 (0.71) | 0.814 | 36.65 (0.71) | 36.70 (0.65) | 0.045 | 36.66(0.68) | 36.74 (0.72) | 0.037 |
| SpO2 (%) (mean (SD)) | 96.76 (3.67) | 95.70 (4.92) | 0.008 | 96.99 (3.50) | 96.20 (4.14) | <0.001 | 96.71 (3.74) | 96.55 (4.00) | 0.459 |
| Urine output (mL) (mean (SD)) | 1802.33 (1333.75) | 1341.22 (981.83) | <0.001 | 1862.26 (1290.47) | 1620.42 (1348.83) | <0.001 | 1792.70 (1325.29) | 1637.70 (1264.41) | 0.047 |
| Creatinine (mg/dL) (mean (SD)) | 1.18 (0.98) | 1.08 (0.67) | 0.070 | 1.12 (0.99) | 1.27 (0.91) | <0.001 | 1.19 (1.00) | 1.06 (0.72) | 0.004 |
| Hemoglobin (gm/dl) (mean (SD)) | 10.18 (2.27) | 9.60 (2.08) | 0.002 | 10.32 (2.33) | 9.85 (2.11) | <0.001 | 10.15 (2.28) | 10.06 (2.15) | 0.494 |
| Myocardial infarction (%) | 887 (40.25) | 62 (38.51) | 0.678 | 588 (39.95) | 361 (40.43) | 0.829 | 806 (39.65) | 143 (43.07) | 0.251 |
| Cerebrovascular disease (%) | 298 (13.52) | 39 (24.22) | <0.001 | 194 (13.18) | 143 (16.01) | 0.060 | 281 (13.82) | 56 (16.87) | 0.150 |
| Chronic pulmonary disease (%) | 714 (32.40) | 53 (32.92) | 0.931 | 435 (29.55) | 332 (37.18) | <0.001 | 650 (31.97) | 117 (35.24) | 0.255 |
| Diabetes mellitus (%) | 1048 (47.55) | 67 (41.61) | 0.164 | 670 (45.52) | 445 (49.83) | 0.046 | 960 (47.22) | 155 (46.69) | 0.859 |
| Renal disease (%) | 965 (43.79) | 81 (50.31) | 0.118 | 564 (38.32) | 482 (53.98) | <0.001 | 892 (43.88) | 154 (46.39) | 0.404 |
| GCS (mean (SD)) | 13.15 (2.85) | 11.32 (3.71) | <0.001 | 13.27 (2.88) | 12.64 (3.04) | <0.001 | 13.08 (2.93) | 12.73 (3.11) | 0.051 |
| SOFA score (mean (SD)) | 5.79 (3.47) | 6.84 (3.48) | <0.001 | 5.62 (3.43) | 6.28 (3.52) | <0.001 | 5.82 (3.44) | 6.14 (3.70) | 0.128 |
| CCI (mean (SD)) | 7.81 (2.55) | 9.25 (2.45) | <0.001 | 7.27 (2.43) | 8.95 (2.45) | <0.001 | 7.83 (2.53) | 8.38 (2.77) | <0.001 |
| APS III (mean (SD)) | 48.10 (18.86) | 60.75 (20.42) | <0.001 | 45.35 (18.24) | 54.92 (19.35) | <0.001 | 48.33 (18.84) | 52.86 (21.06) | <0.001 |
| SAPSII score (mean (SD)) | 38.84 (11.65) | 45.43 (10.35) | <0.001 | 37.13 (11.51) | 42.86 (11.09) | <0.001 | 39.09 (11.56) | 40.55 (12.35) | 0.034 |

Abbreviations: BMI = body mass index, MBP = mean blood pressure, HR = heart rate, aHR = 24-hour averaged heart rate, GCS = Glasgow coma scale, SOFA = sequential organ failure assessment, CCI = Charlson
comorbidity score, APS III = acute physiology score III, SAPSII = simplified acute physiology score II

### Statistical Analysis

Continuous variables are expressed as the mean and standard deviation (SD). Categorical variables are presented as the number and percentage (%). The Chi-square test or Fisher’s exact test was used to test comparisons between groups for categorical variables, whereas the t test was used for comparisons of continuous variables as appropriate. We used receiver operating characteristic curves to identify the last heart rate before leaving the ICU cutoff values for each study outcome. Logistic regression analysis was conducted to assess the association between heart rate and outcomes, with the results expressed as odds ratios (ORs) and corresponding 95% confidence intervals (95% CIs). Kaplan–Meier survival curves were produced for three outcomes: in-hospital mortality, total mortality and ICU readmission. All analyses were performed in R (4.2.3) and SPSS (20). A threshold of p < 0.05 (two-sided) was considered statistically significant.

## Results

### Study Population

Considering the 76943 admissions registered in the ICU, we enrolled 2365 adult HF patients in this study (Table 1). During the follow-up period of 140.26±246.02 days, the average length of ICU stay was 4.49±5.10 days, and the mean duration of total hospital stay was 13.91±12.44 days. The female proportion was 40.47% (n=957), the mean age of the study population was 69.75 ± 12.78 years, and the BMI was 29.93 ± 8.18Kg/m^2^. The mean heart rate during admission to the ICU was 87.66 ± 20.46 bpm, the mean ICU discharged heart rate was 84.26 ± 16.12 bpm, and the mean 24-hour average ICU discharged heart rate was 83.82 ± 14.49 bpm. The underlying diseases of the enrolled population included myocardial infarction (40.1%, n=949), peripheral vascular disease (16.7%, n=396), cerebrovascular disease (14.3%, n=337) and diabetes mellitus (47.2%, n=1115).

### Clinical Characteristics and Outcomes

The in-hospital mortality was 6.8% (n=161), total mortality was 37.8% (n=893) and ICU readmission rate was 14% (n=332). In terms of in-hospital/total mortality and ICU readmission, no significant difference was observed across sexes. Patients who were younger and had a lower body mass index (BMI) demonstrated a lower rate of in-hospital mortality (p = 0.001) and total mortality (p < 0.001) (Table 1). The in-hospital mortality, total mortality and ICU readmission were all related to lower ICU discharged heart rate, 24-hour average ICU discharged heart rate and ICU admission heart rate. Regarding the outcome of in-hospital mortality, the ICU discharged heart rate was 83.56±15.81 bpm (survival) vs 93.84±17.28 bpm (dead) (p<0.001), and the 24-hour average ICU discharged heart rate was 83.20±14.29 bpm (survival) vs 92.41±14.55 bpm (dead) (p<0.001). Regarding the outcome of total mortality, the ICU discharged heart rate was 83.67±15.36 bpm (survival) vs 85.23±17.25 bpm (dead) (p=0.027), and the 24-hour average ICU discharged heart rate was 83.18±14.00 bpm vs 84.89±15.21 bpm (p=0.006). Regarding the outcome of ICU readmission, the ICU discharged heart rate was 83.55±15.77 bpm (not ICU readmission vs 88.64±17.49 bpm (ICU readmission) (p<0.001), and the 24-hour average ICU discharged heart rate was 83.15±14.42 bpm vs 87.94±14.23 bpm (p<0.001) (Table 1). eFigure 1 shows the area under the curve for the study outcomes. According to the receiver operating characteristic (ROC) curve, the optimal cutoff points for in-hospital mortality, total mortality, and ICU readmission were 87.5 bpm, 91.5 bpm, and 89.5 bpm, respectively. Thus, for this study, we selected 90 bpm as the threshold for the correlation with our outcomes of interest.

The primary and secondary outcomes were significantly higher in the group with HR > 90 bpm (eTable 2). The univariate analysis showed that elevated ICU discharge heart rate was associated with significantly increased in-hospital mortality (OR 1.036 [95% CI 1.027L1.046], p

< 0.001), total mortality (OR 1.006 [95% CI 1.001L1.011], p = 0.023) and ICU readmission rate (OR 1.019 [95% CI 1.012L1.026], p < 0.001). After multivariate analysis, the ICU discharge heart rate was also observed to be a strong predictor of in-hospital mortality (OR 1.035 [95% CI 1.024L1.046], p < 0.001), total mortality (OR 1.007 [95% CI 1.001L1.014], p = 0.027) and ICU readmission (OR 1.015 [95% CI 1.007L1.023], p < 0.001). An ICU-discharged heart rate greater than 90 bpm was significantly related to in-hospital mortality (OR 2.986 [95% CI 2.066L4.315], p < 0.001), total mortality (OR 1.341 [95% CI 1.083L1.661], p = 0.007), and ICU readmission (OR 1.638 [95% CI 1.270L2.114], p < 0.001) (Table 2). The KaplanLMeier analysis also showed that an HR > 90 bpm was significantly related to in-hospital mortality (p<0.001), total mortality (p<0.001) and ICU readmission (p<0.001) (Figure 2).

**Figure 2.**
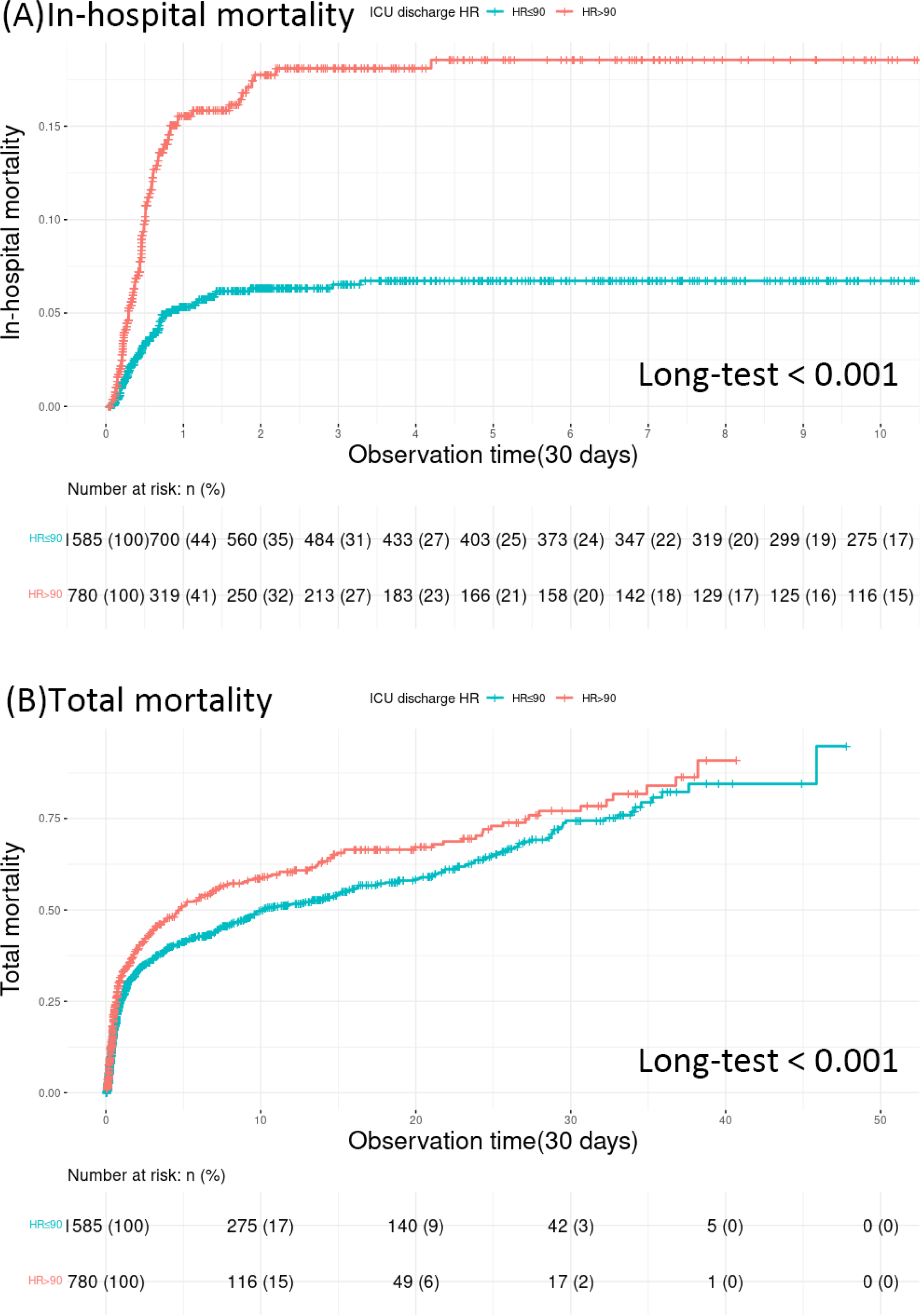
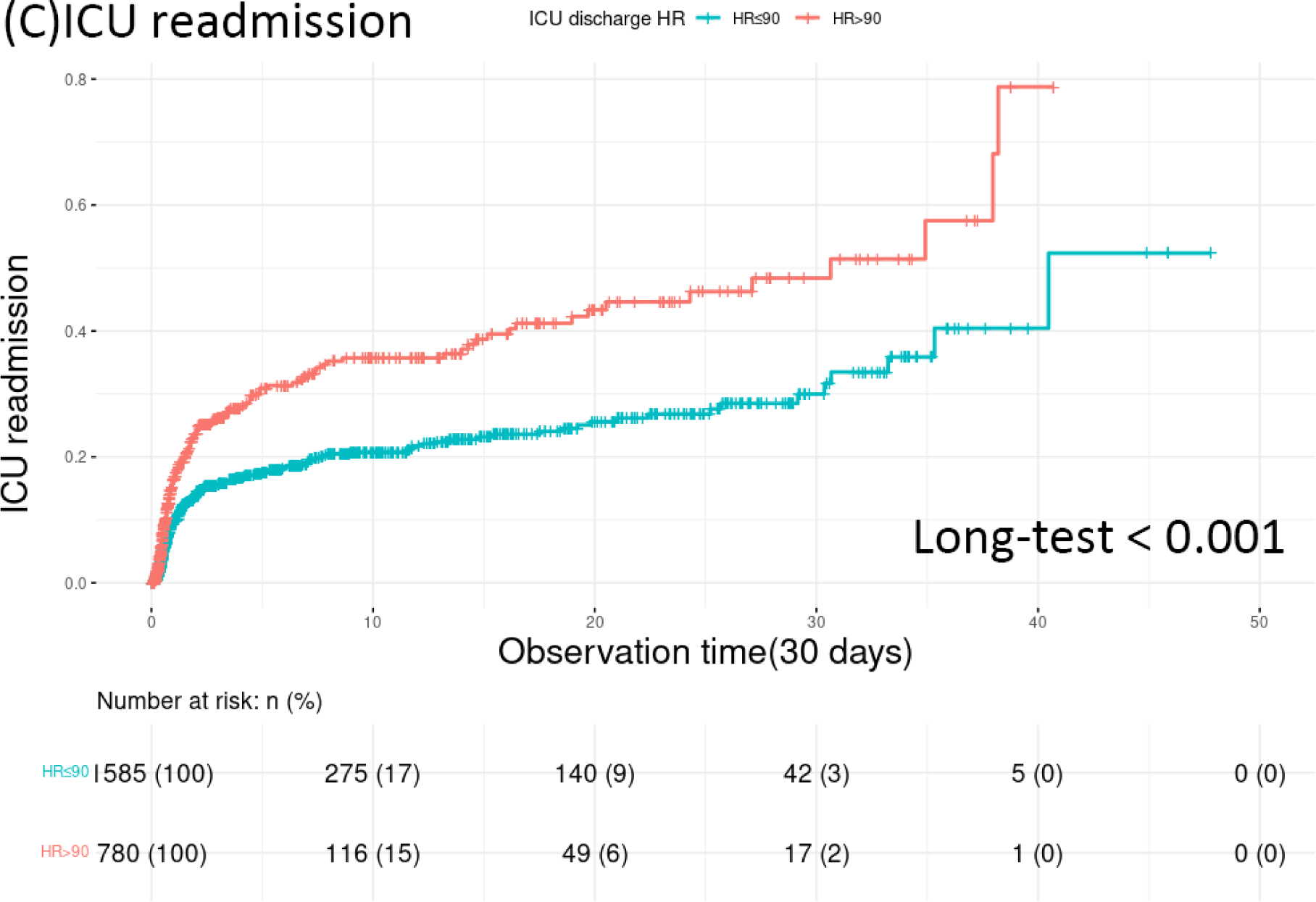
KaplanLMeier curves show that the risks of follow-up all-cause (A), in-hospital mortality (B), total mortality (C), and ICU readmission were significantly higher in HF patients with heart rate > 90 bpm than in those with heart rate ≤ 90bpm

**Table 2.** Variables associated with study outcomes in logistic regression analyses.

| Variable | cOR | (95% CI) |  | P value | aOR | (95% CI) |  | P value | aOR | (95% CI) |  | P value |
| --- | --- | --- | --- | --- | --- | --- | --- | --- | --- | --- | --- | --- |
| In-hospital mortality |  |  |  |  |  |  |  |  |  |  |  |  |
| ICU discharge HR as a continuous variable<sup>a</sup> | 1.036 | 1.027 | 1.046 | <0.001 | 1.035 | 1.024 | 1.046 | <0.001 |  |  |  |  |
| ICU discharge HR > 90 bpm vs. HR ≤ 90 bpm<sup>a</sup> | 3.192 | 2.302 | 4.425 | <0.001 |  |  |  |  | 2.986 | 2.066 | 4.315 | <0.001 |
| ICU discharge aHR | 1.042 | 1.031 | 1.053 | <0.001 |  |  |  |  |  |  |  |  |
| ICU admission HR | 1.017 | 1.010 | 1.024 | <0.001 | 1.006 | 0.996 | 1.015 | 0.244 | 1.008 | 0.999 | 1.017 | 0.091 |
| Female | 1.142 | 0.827 | 1.578 | 0.420 | 0.890 | 0.622 | 1.274 | 0.525 | 0.879 | 0.615 | 1.257 | 0.480 |
| Age | 1.042 | 1.027 | 1.057 | <0.001 | 1.032 | 1.015 | 1.049 | <0.001 | 1.032 | 1.015 | 1.049 | <0.001 |
| BMI | 0.959 | 0.936 | 0.982 | 0.001 | 0.977 | 0.951 | 1.003 | 0.078 | 0.977 | 0.952 | 1.003 | 0.085 |
| MBP | 1.001 | 0.993 | 1.009 | 0.857 |  |  |  |  |  |  |  |  |
| Urine output | 1.000 | 0.999 | 1.000 | <0.001 | 1.000 | 1.000 | 1.000 | 0.009 | 1.000 | 1.000 | 1.000 | 0.011 |
| Creatinine | 0.870 | 0.706 | 1.071 | 0.188 |  |  |  |  |  |  |  |  |
| Hemoglobin | 0.889 | 0.825 | 0.957 | 0.002 | 0.876 | 0.804 | 0.954 | 0.002 | 0.889 | 0.816 | 0.968 | 0.007 |
| Myocardial infarction | 0.930 | 0.669 | 1.292 | 0.665 |  |  |  |  |  |  |  |  |
| Cerebrovascular disease | 2.045 | 1.397 | 2.992 | <0.001 | 1.386 | 0.894 | 2.149 | 0.145 | 1.447 | 0.934 | 2.242 | 0.098 |
| Chronic pulmonary disease | 1.024 | 0.728 | 1.440 | 0.891 |  |  |  |  |  |  |  |  |
| Diabetes | 0.786 | 0.568 | 1.087 | 0.146 |  |  |  |  |  |  |  |  |
| Renal disease | 1.300 | 0.944 | 1.791 | 0.108 |  |  |  |  |  |  |  |  |
| GCS | 0.861 | 0.827 | 0.897 | <0.001 | 0.869 | 0.809 | 0.934 | <0.001 | 0.869 | 0.809 | 0.933 | <0.001 |
| SOFA score | 1.084 | 1.038 | 1.131 | <0.001 | 0.981 | 0.917 | 1.050 | 0.579 | 0.980 | 0.915 | 1.049 | 0.554 |
| CCI | 1.227 | 1.156 | 1.302 | <0.001 | 1.160 | 1.081 | 1.245 | <0.001 | 1.151 | 1.073 | 1.236 | <0.001 |
| APS III | 1.028 | 1.021 | 1.035 | <0.001 | 1.005 | 0.991 | 1.020 | 0.501 | 1.006 | 0.992 | 1.021 | 0.388 |
| Total mortality |  |  |  |  |  |  |  |  |  |  |  |  |
| ICU discharge HR as a continuous variable<sup>a</sup> | 1.006 | 1.001 | 1.011 | 0.023 | 1.007 | 1.001 | 1.014 | 0.027 |  |  |  |  |
| ICU discharge HR > 90 bpm vs. HR ≤ 90 bpm<sup>a</sup> | 1.321 | 1.108 | 1.574 | 0.002 |  |  |  |  | 1.341 | 1.083 | 1.661 | 0.007 |
| ICU discharge aHR | 1.008 | 1.002 | 1.014 | 0.005 |  |  |  |  |  |  |  |  |
| ICU admission HR | 1.005 | 1.001 | 1.009 | 0.014 | 0.997 | 0.992 | 1.002 | 0.257 | 0.997 | 0.992 | 1.002 | 0.253 |
| Female | 1.166 | 0.985 | 1.381 | 0.074 | 0.986 | 0.806 | 1.206 | 0.892 | 0.984 | 0.805 | 1.204 | 0.877 |
| Age | 1.042 | 1.035 | 1.050 | <0.001 | 1.028 | 1.018 | 1.037 | <0.001 | 1.028 | 1.018 | 1.037 | <0.001 |
| BMI | 0.965 | 0.954 | 0.976 | <0.001 | 0.975 | 0.962 | <0.001 | <0.001 | 0.975 | 0.962 | 0.988 | <0.001 |
| MBP | 0.998 | 0.993 | 1.002 | 0.331 |  |  |  |  |  |  |  |  |
| Urine output | 1.000 | 1.000 | 1.000 | <0.001 | 1.000 | 1.000 | 1.000 | 0.641 | 1.000 | 1.000 | 1.000 | 0.644 |
| Creatinine | 1.171 | 1.072 | 1.278 | <0.001 | 1.004 | 0.889 | 1.134 | 0.951 | 1.003 | 0.888 | 1.134 | 0.956 |
| Hemoglobin | 0.911 | 0.877 | 0.946 | <0.001 | 0.943 | 0.901 | 0.986 | 0.011 | 0.944 | 0.902 | 0.988 | 0.013 |
| Myocardial infarction | 1.020 | 0.861 | 1.209 | 0.817 |  |  |  |  |  |  |  |  |
| Cerebrovascular disease | 1.256 | 0.994 | 1.587 | 0.056 |  |  |  |  |  |  |  |  |
| Chronic pulmonary disease | 1.411 | 1.183 | 1.682 | <0.001 | 1.195 | 0.970 | 1.472 | 0.094 | 1.186 | 0.963 | 1.462 | 0.108 |
| Diabetes | 1.189 | 1.007 | 1.404 | 0.042 | 0.838 | 0.673 | 1.044 | 0.115 | 0.834 | 0.670 | 1.039 | 0.106 |
| Renal disease | 1.888 | 1.595 | 2.234 | <0.001 | 0.767 | 0.591 | 0.996 | 0.046 | 0.762 | 0.587 | 0.989 | 0.041 |
| GCS | 0.932 | 0.907 | 0.958 | <0.001 | 1.017 | 0.973 | 1.064 | 0.446 | 1.017 | 0.973 | 0.458 |  |
| SOFA score | 1.056 | 1.031 | 1.081 | <0.001 | 0.988 | 0.950 | 1.027 | 0.530 | 0.988 | 0.950 | 0.528 |  |
| CCI | 1.320 | 1.271 | 1.370 | <0.001 | 1.274 | 1.206 | 1.346 | <0.001 | 1.275 | 1.207 | <0.001 |  |
| APS III | 1.027 | 1.022 | 1.032 | <0.001 | 1.028 | 1.019 | 1.036 | <0.001 | 1.028 | 1.019 | <0.001 |  |
| ICU readmission |  |  |  |  |  |  |  |  |  |  |  |  |
| ICU discharge HR as a continuous variable<sup>a</sup> | 1.019 | 1.012 | 1.026 | <0.001 | 1.015 | 1.007 | 1.023 | <0.001 |  |  |  |  |
| ICU discharge HR > 90 bpm vs. HR ≤ 90 bpm<sup>a</sup> | 1.862 | 1.471 | 2.357 | <0.001 |  |  |  |  | 1.638 | 1.270 | 2.114 | <0.001 |
| ICU discharge aHR | 1.022 | 1.014 | 1.031 | <0.001 |  |  |  |  |  |  |  |  |
| ICU admission HR | 1.009 | 1.004 | 1.015 | 0.001 | 1.000 | 0.993 | 1.006 | 0.901 | 1.000 | 0.994 | 1.007 | 0.905 |
| Female | 1.010 | 0.797 | 1.279 | 0.937 | 0.871 | 0.678 | 1.118 | 0.278 | 0.862 | 0.671 | 1.107 | 0.244 |
| Age | 0.998 | 0.989 | 1.007 | 0.618 | 0.990 | 0.980 | 1.000 | 0.050 | 0.990 | 0.980 | 1.000 | 0.056 |
| BMI | 0.997 | 0.983 | 1.011 | 0.640 | 0.994 | 0.980 | 1.009 | 0.465 | 0.995 | 0.980 | 1.010 | 0.473 |
| MBP | 1.002 | 0.996 | 1.007 | 0.622 |  |  |  |  |  |  |  |  |
| Urine output | 1.000 | 1.000 | 1.000 | 0.047 | 1.000 | 1.000 | 1.000 | 0.044 | 1.000 | 1.000 | 1.000 | 0.044 |
| Creatinine | 0.835 | 0.715 | 0.975 | 0.022 | 0.728 | 0.608 | 0.873 | 0.001 | 0.720 | 0.601 | 0.863 | <0.001 |
| Hemoglobin | 0.982 | 0.933 | 1.034 | 0.494 |  |  |  |  |  |  |  |  |
| Myocardial infarction | 1.152 | 0.911 | 1.457 | 0.238 |  |  |  |  |  |  |  |  |
| Cerebrovascular disease | 1.265 | 0.924 | 1.731 | 0.142 |  |  |  |  |  |  |  |  |
| Chronic pulmonary disease | 1.158 | 0.907 | 1.477 | 0.238 |  |  |  |  |  |  |  |  |
| Diabetes | 0.979 | 0.776 | 1.235 | 0.857 |  |  |  |  |  |  |  |  |
| Renal disease | 1.107 | 0.877 | 1.397 | 0.393 |  |  |  |  |  |  |  |  |
| GCS | 0.964 | 0.930 | 1.000 | 0.052 |  |  |  |  |  |  |  |  |
| SOFA score | 1.026 | 0.993 | 1.060 | 0.128 |  |  |  |  |  |  |  |  |
| CCI | 1.085 | 1.038 | 1.134 | <0.001 | 1.107 | 1.054 | 1.162 | <0.001 | 1.105 | 1.052 | 1.160 | <0.001 |
| APS III | 1.011 | 1.006 | 1.017 | <0.001 | 1.008 | 1.002 | 1.015 | 0.009 | 1.009 | 1.002 | 1.015 | 0.007 |

<sup>a</sup> ICU discharge HR as a continuous variable and ICU discharge HR > 90 bpm vs. HR ≤ 90 bpm as a binary variable were not simultaneously adjusted in multivariate logistic regression analyses. cOR = crude odds
ratio; aOR = adjusted odds ratio; CI: confidence interval

## Discussion

In this clinical investigation, we were able to demonstrate that heart rate at ICU discharge was a predictor of mortality in HF patients in the MIMIC IV group. Consistent with prior research on the general population or specific patient groups, we found that a higher heart rate before ICU discharge was associated with increased in-hospital mortality among ICU patients. This association was statistically significant with an adjusted odds ratio (OR) of 1.035 for each 1 bpm increase in heart rate in the ICU. A similar study in Japan reported adjusted ORs of 1.03 and 1.02 for each 1 bpm increment in HR in medical and surgical wards, respectively (19). A study has shown that heart rate upon admission can predict in-hospital mortality in patients with ischemic stroke, with a hazard ratio of 4.42 for a 10 bpm increase in heart rate (20). Furthermore, high heart rate 24-36 hours after admission has also been associated with in-hospital mortality in patients with HF (21).

A study found that keeping heart rate below 100 bpm within the first day of admission could decrease mortality in ICU patients (22). Another study showed that heart rate measured 24 hours before ICU discharge was independently linked to in-hospital and posthospital mortality after ICU discharge (23). One study using the MIMIC-III database showed a significant association of prolonged elevated heart rate with decreased survival in a large and heterogeneous cohort of ICU patients (24). A separate study utilizing the MIMIC-IV database demonstrated that the group of patients with rheumatic heart disease who survived had a lower heart rate (25). The HF mortality prediction models have served to advance different perspectives on whether heart rate is a predictor or not. Some models suggest a strong impact on prognosis, such as the OPTIMIZE-HF study and another study that uses a GWTG-HF risk score (8, 9, 26) and both utilized heart rate at admission as a predictor. To our knowledge, there has been no report linking ICU-discharge heart rate and mortality among HF patients in the ICU. Our study is the first to explore this important clinical issue. In our study, a higher last heart rate before ICU discharge was significantly associated with in-hospital/total mortality and ICU readmission. Heart rate greater than and equal to 90 bpm was strongly associated with increased in-hospital/total mortality and ICU readmission rates.

It is well established that there is a link between heart rate and mortality. Multiple theories have been proposed to explain the association, including low physical fitness, elevated blood pressure, reduced variability in heart rate, and decreased baroreceptor sensitivity in individuals with high heart rate (27, 28). However, the underlying causes of this correlation have yet to be fully understood (29). The significance of heart rate as a prognostic indicator in ADHF remains a topic of debate. Unlike its predictive role in chronic systolic HF, the role of heart rate in ADHF is much less clear. This is partly due to variations in the timing of heart rate measurement during an acute episode and differences in the outcome measures used in various studies, such as in-hospital mortality and readmission (30). When patients with ADHF presented at the emergency department with a higher heart rate, a higher risk of death within 7 days was linked (31). Patients with ADHF who had a higher heart rate upon arriving at the hospital were more likely to have a higher risk of death while in the hospital, with the lowest risk observed at heart rates between 70-75 bpm (32).

The SHIFT trial has proved that heart rate plays a role in the progression of HF (12), according the speculation about a fundamental connection between a higher heart rate and the development (33) or worsening of HF (10), with theories ranging from the impact on myocardial energy (34) to the potential benefits of reducing arterial afterload by reducing heart rate (35).

Our findings are consistent with a large retrospective cohort study that examined HF patients who had been treated for ADHF. This study found that a higher heart rate at discharge after treatment was linked to a higher risk of death and rehospitalization, with an even greater risk within the first 30 days following discharge (36). Using heart rate at discharge as a predictor of risk has several benefits, including being routinely available, being easy and reliable to determine, and reflective of the patient’s most stable condition, which may be more relevant for future prognosis.

Despite the large sample size and detailed data, this study has several limitations that are common to all retrospective cohort studies. These include missing data, the potential for bias in the data collection and analysis, and the inability to account for all possible confounding factors. Given that the data we used originated from an observational database, we were unable to analyze specific causes of death and could only evaluate the overall mortality rate. We were unable to conduct an analysis of major adverse cardiovascular events (MACEs) due to this limitation. Furthermore, we lacked detailed information on echocardiography, specific therapeutic medications, and whether cardiac implantable electronic devices (CIEDs) were installed. Therefore, the results reported in our study should be regarded only as reference points and must be further verified through additional research.

## Conclusions

Given the association between elevated heart rate at ICU discharge in patients with HF and increasing in-hospital mortality, total mortality, and ICU readmission, it is evident that a heart rate exceeding 90 bpm correlates with an increase in these outcomes. It is important to conduct further research to determine if reducing heart rate could be an effective treatment strategy for HF patients discharged from the ICU.

## Data Availability

All data produced in the present study are available upon reasonable request to the authors

## Acknowledgments

Special thanks to Lang-Yin Zeng for her efforts on this article.

