## Supplement Table 1 for "Heart Rate as a Predictor of Mortality in Heart Failure Patients at the Time of Discharge from the Intensive Care Unit"

**eTable 1.** Baseline characteristics

| Variable | All patients, n=2365 |
| --- | --- |
| Sex (Female) (%) | 957(40.47) |
| Age (mean (SD)) | 69.7(12.69) |
| Weight (kg) (mean (SD)) | 84.86(25.34) |
| Height (cm) (mean (SD)) | 168.45(10.97) |
| BMI (kg/m^2^) (mean (SD)) | 29.83(8.31) |
| MBP (mmHg) (mean (SD)) | 84.13(19.15) |
| ICU Discharge HR (bpm) (mean (SD)) | 84.26 (16.12) |
| ICU Discharge aHR (bpm) (mean (SD)) | 83.82(14.49) |
| ICU admission HR (bpm) (mean (SD)) | 87.66(20.46) |
| Temperature (°C) (mean (SD)) | 36.67(0.69) |
| SpO2(%) (mean (SD)) | 96.69(3.77) |
| Urine output (mL) (mean (SD)) | 1770.94(1317.76) |
| Creatinine (mg/dL) (mean (SD)) | 1.17(0.96) |
| Hemoglobin (gm/dl) (mean (SD)) | 10.14(2.26) |
| Myocardial infarction (%) | 949(40.13) |
| Cerebrovascular disease (%) | 337(14.25) |
| Chronic pulmonary disease (%) | 767(32.43) |
| Diabetes mellitus (%) | 1115(47.15) |
| Renal disease (%) | 1046(44.23) |
| GCS (mean (SD)) | 13.03(2.96) |
| SOFA score (mean (SD)) | 5.87(3.48) |
| CCI (mean (SD)) | 7.90(2.57) |
| APS III (mean (SD)) | 48.96(19.23) |
| SAPSII score (mean (SD)) | 39.29(11.69) |
| Follow-up period (day) (mean (SD)) | 140.26(246.02) |
| Length of ICU stay (day) (mean (SD)) | 4.49(5.10) |
| Length of hospital stay (day) (mean (SD)) | 13.91(12.44) |
| In-hospital mortality (%) | 161(6.81) |
| Total mortality (%) | 893(37.76) |
| ICU readmission (%) | 332(14.04) |

Abbreviation: BMI = body mass index, MBP = mean blood pressure, HR = heart rate, aHR = 24-hour averaged heart rate, GCS = Glasgow coma scale, SOFA = sequential organ failure assessment, CCI = Charlson comorbidity score, APS III = acute physiology score III, SAPSII = simplified acute physiology score II
