## Supplement Table 2 for "Heart Rate as a Predictor of Mortality in Heart Failure Patients at the Time of Discharge from the Intensive Care Unit"

**eTable 2.** Outcome analysis by HR > 90 bpm vs. HR ≤ 90 bpm

|  | In-hospital mortality | | | | Total mortality | | | | ICU readmission | | | |
| --- | --- | --- | --- | --- | --- | --- | --- | --- | --- | --- | --- | --- |
| HR | Survival | Dead | Total | *p* | Survival | Dead | Total | *p* | No | Yes | Total | *p* |
| HR > 90 | 1519 (95.84) | 66 (4.16) | 1585 | <0.001 | 1021 (64.42) | 564 (35.58) | 158 | 0.002 | 1404 (88.58) | 181 (11.42) | 1585 | <0.001 |
| HR $\leq$ 90 | 685 (87.82) | 95 (12.18) | 780 |  | 451 (57.82) | 329 (42.18) | 780 |  | 629 (80.64) | 151 (19.36) | 780 |  |
| Total | 2204 (93.19) | 161 (6.81) |  |  | 1472 (62.24) | 893 (37.76) |  |  | 2033 (85.96) | 332 (14.04) |  |  |
