## Supplement Figure 1 for "Heart Rate as a Predictor of Mortality in Heart Failure Patients at the Time of Discharge from the Intensive Care Unit"

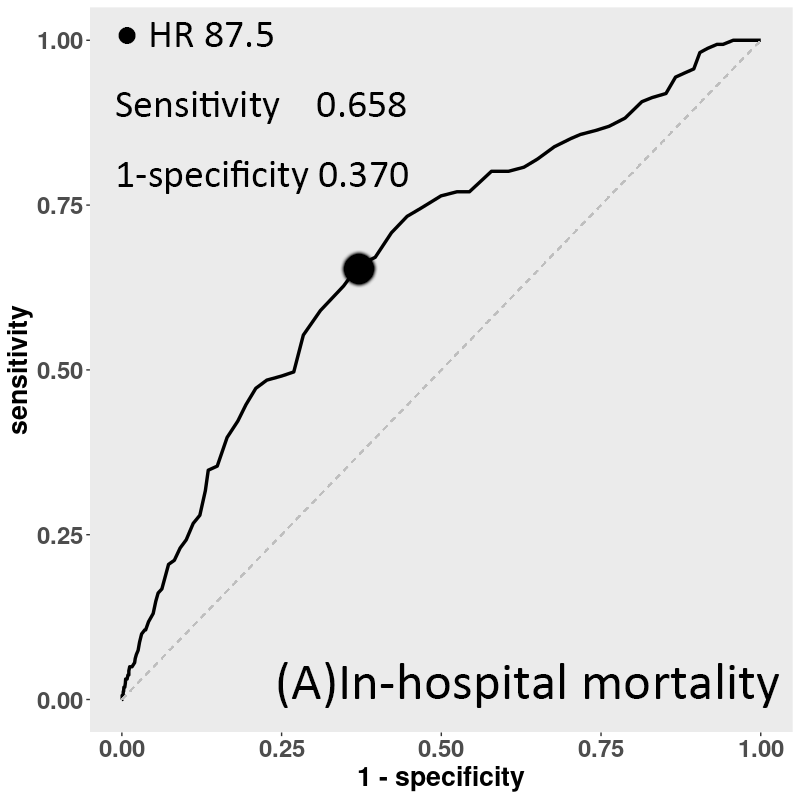

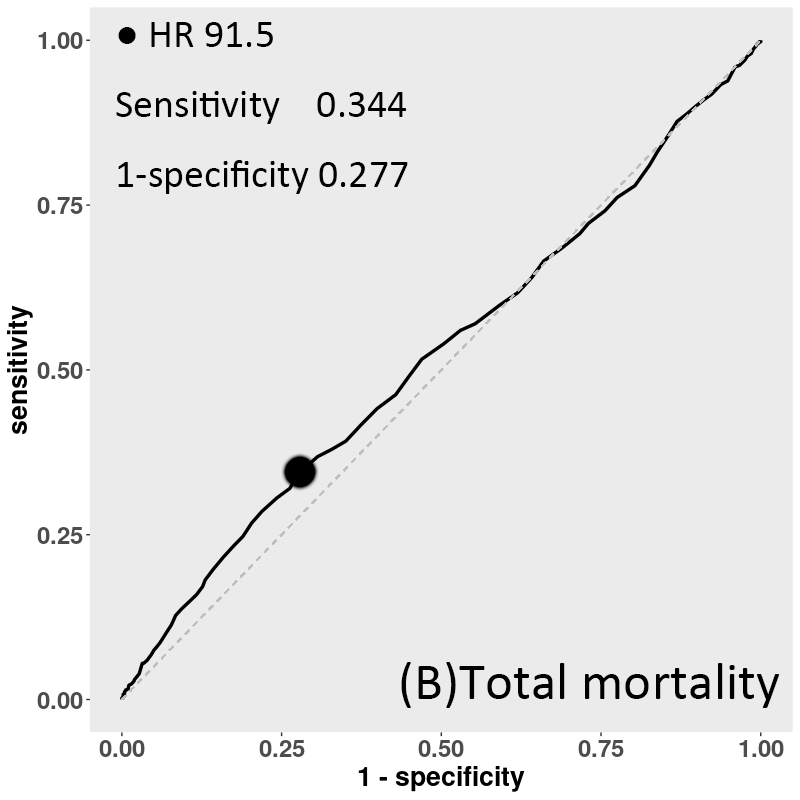

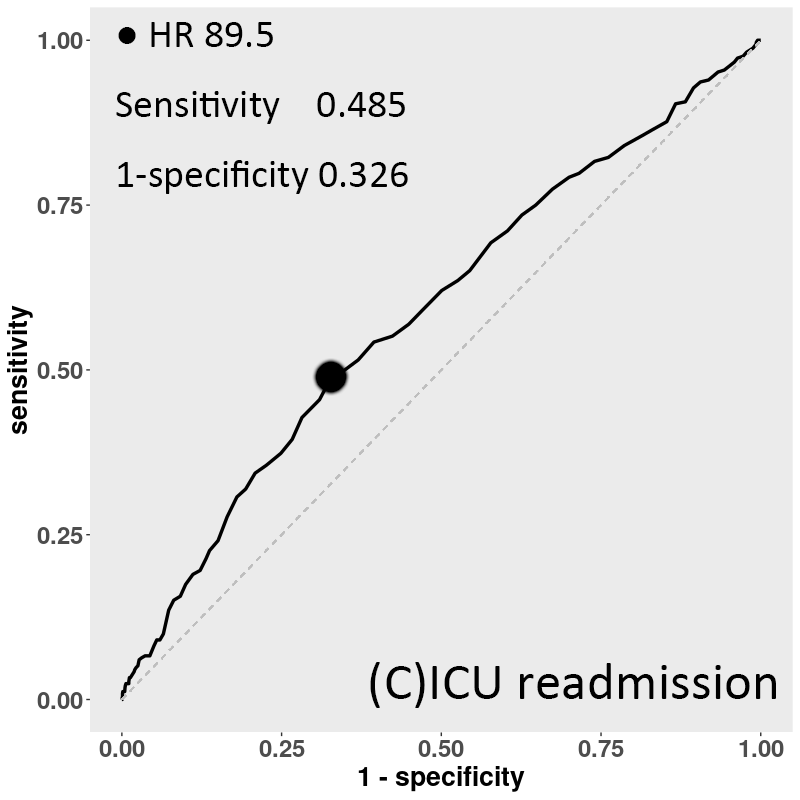

**eFigure 1.** Receiver operating characteristic curves showing the ICU discharge HR cutoff for each study outcome. (A), In-hospital mortality (B), total mortality (C) and ICU readmission
